# Pre-dominance of dengue non-cross-reacting SARS-CoV-2 spike antibodies during the Omicron era and their role in the ADE-mediated surge of Dengue virus serotype 3

**DOI:** 10.1101/2023.09.08.23295136

**Authors:** Supratim Sarker, Chiroshri Dutta, Abinash Mallick, Sayantan Das, Chandrika Das Chowdhury, Abhishek De, Surajit Gorai, Subhajit Biswas

**Affiliations:** Infectious Diseases and Immunology Division, CSIR-Indian Institute of Chemical Biology, West Bengal, India; Department of Dermatology, Calcutta National Medical College and Hospital, Kolkata, West Bengal, India; Department of Dermatology, Apollo Multispeciality Hospital, Kolkata, West Bengal, India; Academy of Scientific and Innovative Research (AcSIR), Ghaziabad-201002, India

**Keywords:** Dengue, SARS-CoV-2, cross-reactivity, ADE, dengue serotype 3, virus neutralization test

## Abstract

In India, the SARS-CoV-2 Delta wave (2020-21) gradually faded away with the advent of the Omicron variants (2021-present). Dengue incidences were observably less in Southeast Asia during the active years of the pandemic (2020-21). However, Dengue virus type 3 (DV3) cases were increasingly reported in India and many other dengue-endemic countries concurrent with the progression of the Omicron wave since 2022. This observation prompted us to investigate the current state of cross-reactivity between prevalent SARS-CoV-2 variants and different DV serotypes.

Fifty-five COVID-19 serum samples (collected between January-September, 2022) and three pre-pandemic healthy serum samples were tested for DV or SARS-CoV-2 Immunoglobulin G/Immunoglobulin M (IgG/IgM) using the lateral flow immunoassays (LFIAs). The SARS-CoV-2 antibody (Ab)-positive samples were further tested for their ability to cross-neutralize DV types 1-4 (DV1-4) in Huh7 cell lines.

Cross-reactivity between SARS-CoV-2 and DV diminished with the shift from Delta to Omicron prevalence. COVID-19 serum samples that were DV cross-reactive neutralized all DV serotypes, including DV3. However, Omicron wave serum samples were predominantly DV non-cross-reactive (about 70%) in LFIAs and coincided with the prevalence of BA.2 Omicron variant. They also cross-neutralized DV1, 2 and 4 but enhanced DV3 infectivity as evident from increased DV3 titres in virus neutralization test (VNT) compared to control serum samples.

In conclusion, DV non-cross-reactive COVID-19 serum samples are becoming increasingly prominent in the present times. These non-cross-reactive serums could neutralize DV1, 2 and 4 but they are contributing to the surge in DV3 cases worldwide by means of ADE.

## 1. INTRODUCTION

SARS-CoV-2 coronavirus (Family: *Coronaviridae*) is the cause of the deadly and contagious COVID-19 pandemic (acute respiratory illness). COVID-19 has posed a major hazard to public health worldwide^1^. We had observed that SARS-CoV-2 showed a trend of decreased transmission, severity and overall fatality per million populations in the highly dengue-endemic regions^2, 3^. Computational modeling studies from our laboratory supported the hypothesis that dengue virus (DV) (Family: *Flaviviridae*) envelope antibodies (Abs) can bind to SARS-CoV-2 receptor binding sites^4^. Sero-diagnostic tests for SARS-CoV-2 have also been found to frequently produce false-positive results for pre-pandemic dengue serum samples and *vice versa* in dengue-endemic areas according to several investigations^5, 6^.

Conversely, docking studies revealed that SARS-CoV-2 spike Abs could also significantly bind to DV2 envelope^7^. It is equally interesting to note that during the COVID-19 Delta wave (September, 2020-January, 2021), India had a sharp decline in the number of DV cases^8, 9^. Our previous study revealed that almost 93% of the SARS-CoV-2 Ab-positive serum samples (mostly Delta), collected during the above period, cross-reacted with DV in LFIA or ELISA tests^7^. About 57% of these SARS-CoV-2 serum samples had no evidence of DV pre-exposure (DV-NS1 Ab-negative) and could still “neutralize” the DV1 serotype in virus neutralization test (VNT)^7^. Antibodies raised against the receptor-binding domain (RBD) of S1 subunit of SARS-CoV-2 spike protein and serum samples from COVID-19 patients were shown to neutralize DV2 in a subsequent investigation from Taiwan^10^. All these studies confirmed that there is indeed, an antigenic relationship between SARS-COV-2 and DV and the two viruses are co-evolving under self and mutually cross-reactive immune selection pressures in regions where they are co-endemic now^3^.

In recent times (2022-), there has been an increase in DV3 cases in India and many other dengue endemic regions globally^11–16^ (DV prevalence, Supplementary Table S1). For instance, Kerala, a south-Indian state reported a surge in DV3 cases in 2022, a shift from the previously dominant DV2 serotype^17^. Again, Singapore reported large numbers of dengue cases mainly due to DV3, previously uncommon to that region^18^. We therefore, investigated whether the rapidly emerging Omicron variants have the same or different cross-reactivity/cross-protection pattern against DV serotypes.

## 2. MATERIALS AND METHODS

### Study subjects

55 clinically and laboratory-confirmed (swab reverse transcription-polymerase chain reaction [RT-PCR]-positive) COVID-19 patients’ serum samples (Patient 1-55, Supplementary Tables S2-S4) were collected from Apollo Multispecialty Hospital, Kolkata, from January to September, 2022. All COVID-19 patients showed mild to severe COVID-19 symptoms but were discharged from the hospital eventually on recovery. The study was approved by the respective Institutional Ethical Committees of the previously mentioned hospital and the Council of Scientific & Industrial Research-Indian Institute of Chemical Biology (CSIR-IICB), Kolkata. Written informed consent (in their native language) was obtained from all individual participants included in this study. All experiments were carried out as per relevant guidelines and regulations.

### Dengue and COVID-19 LFIAs

DV-specific IgG, IgM, and NS1 Ag (antigen) detection was done using the Standard Diagnostics-Bioline Dengue Duo rapid test kit while COVID-19-specific IgG and IgM detection was carried out using the Abcheck kit as described in a previous study^7^. All tests were done per the manufacturer’s instructions.

### The NS1 Ab ELISA

The NS1 Ab ELISA was carried out according to the manufacturer’s instructions (R&D Systems, Cat-DENG00), as was performed in our earlier study^7^.

### RT-PCR for DV

RNA extraction was done from 200 µl of COVID-19/healthy serum samples using the High Pure Viral Nucleic Acid Extraction Kit (Roche) as per the manufacturer’s protocol. RT-PCR was done to detect the presence of DV RNA using primers, as described by Lanciotti *et al.*^19^.

### Cell line

Huh7 cells were obtained from National Centre for Cell Science, India. Huh7 cells were cultured in DMEM (Sigma) supplemented with 10% FBS (Gibco), Pen-Strep and L-Glutamine mix (Sigma) and Amphotericin B at 2.5 µg/ml (Gibco). Huh7 cell monolayers were grown at 37°C with 5% CO_2_. During the passage, cells were washed with phosphate-buffered saline (PBS) (1X) and detached with Trypsin-ethylenediaminetetraacetic acid (1X) (Gibco).

### Virus

DV1, DV2 and DV3 were derived from serum samples collected during a dengue outbreak in Kolkata in 2017, as explained previously^20^. The NS1 gene of all three viruses was sequenced and made available in the Genetic Codes Databank (GenBank) under the accession numbers MT072226 (DV1), MT072227 (DV2) and MT072228 (DV3). The passage number was kept low in order to maintain a close resemblance to the clinical conditions. It should be noted that these viruses do not form plaques, similar to other low-passage clinical isolates previously reported^21^. DV4 has been obtained from American Type Culture Collection (ATCC VR1490) and passaged in the Huh7 cell line.

### DV neutralization assay (VNT)

Based on the outcomes of LFIA and ELISA, serum samples were chosen for VNT. The three samples (HS1-3), used as healthy controls for neutralization were archived pre-pandemic serums collected from apparently healthy human beings. These were tested free of DV Abs, SARS-CoV-2 Abs, DV RNA and DV NS1. DV1, DV2 and DV3, without any serum were also included in the study as virus controls. For virus control groups, the viruses were diluted with an equal volume of DMEM1 (1:1). DV neutralizing activity was assessed using both DV cross-reactive and DV non-cross-reactive COVID-19 serum samples in 96-well plates. An equal volume of DMEM (supplemented with Pen-Strep, L-Glutamine mix from Sigma, and Amphotericin B, 2.5 µg/ml) was used to dilute the serum samples after they had been inactivated at 56°C for 45 minutes. 50 µl of diluted (1:1) serum was added to each well of 96-well plates. 50 µl of DV inoculum (100 times the median tissue culture infectious dose of 5 x 10^4^ copies) was then added^22^. Diluted serum samples and DV inoculum were incubated at 37°C with 5% CO_2_ for one hour. Then, 50 µl of Huh7 cell suspension (10^4^ cells) was added to each well, and the plate was kept in an incubator. After 12 hours of incubation, 50 µl of DMEM1 was added to each well. At 96 hours after incubation, the supernatant was aspirated from each well, and cells were washed with 1X PBS twice. After washing, 200 µl of fresh DMEM was added to each well, and cells were harvested; stored at -80°C until RNA was extracted.

### RNA extraction and intracellular virus quantification

RNA extraction was done from 200ul of Huh7 cells and qRT-PCR was done according to our previously described protocol^7^.

### DV4 plaque neutralization assay

2.0 x 10^5^ Huh7 cells were seeded in each well of a 12-well plate. 24 hours after seeding the cells, serum samples were diluted with an equal volume of DMEM1 and incubated at 56°C for 45 minutes. The samples were then incubated at room temperature for 5 minutes. 100 µl (eg. 120 PFU/ml) of DV4 was added to each tube and incubated on a rotor for 1 hour at room temperature. The media was discarded from each well of the 12-well plate and washed with 500 µl of 1XPBS. The mixture (serum plus virus) was added over the monolayer of cells and incubated for 2 hours at 37°C. Intermittent shaking was done every 15 minutes. 2 ml of DMEM1-overlay, thickened with high-density carboxymethylcellulose (CMC) (SIGMA), was added to each of the wells and the plate was incubated at 37°C for 120 hours. After that, the supernatant was discarded from all the wells and 1 ml of crystal violet solution was added to each well and kept for 10 minutes at room temperature. The wells were then washed with water; left overnight for drying and plaque counting was done under microscope.

## 3. RESULTS

### 3.1. Declining cross-reactivity between SARS-CoV-2 and DV with the onset and progression of the Omicron variants in India

3.1.1. **DV cross-reactivity status of COVID-19 serum samples collected during January, 2022**

10 of 13 samples (77%) (collected in January, 2022) were SARS-CoV-2 IgG/IgM/both positive in the COVID-19 strip-based antibody test (LFIA). Again, of these 13 samples, 10 samples (77%) were DV IgG/IgM/both positive in the Dengue Duo strip test. Among the 10 SARS-CoV-2 Ab-positive samples, 8 samples were cross-reactive (80%) in the DV LFIA and 5 of these 8 samples (63%) had no evidence of DV pre-exposure (NS1 Ab-negative). All the samples were DV NS1 antigen negative (Table 1).

**Table 1.**
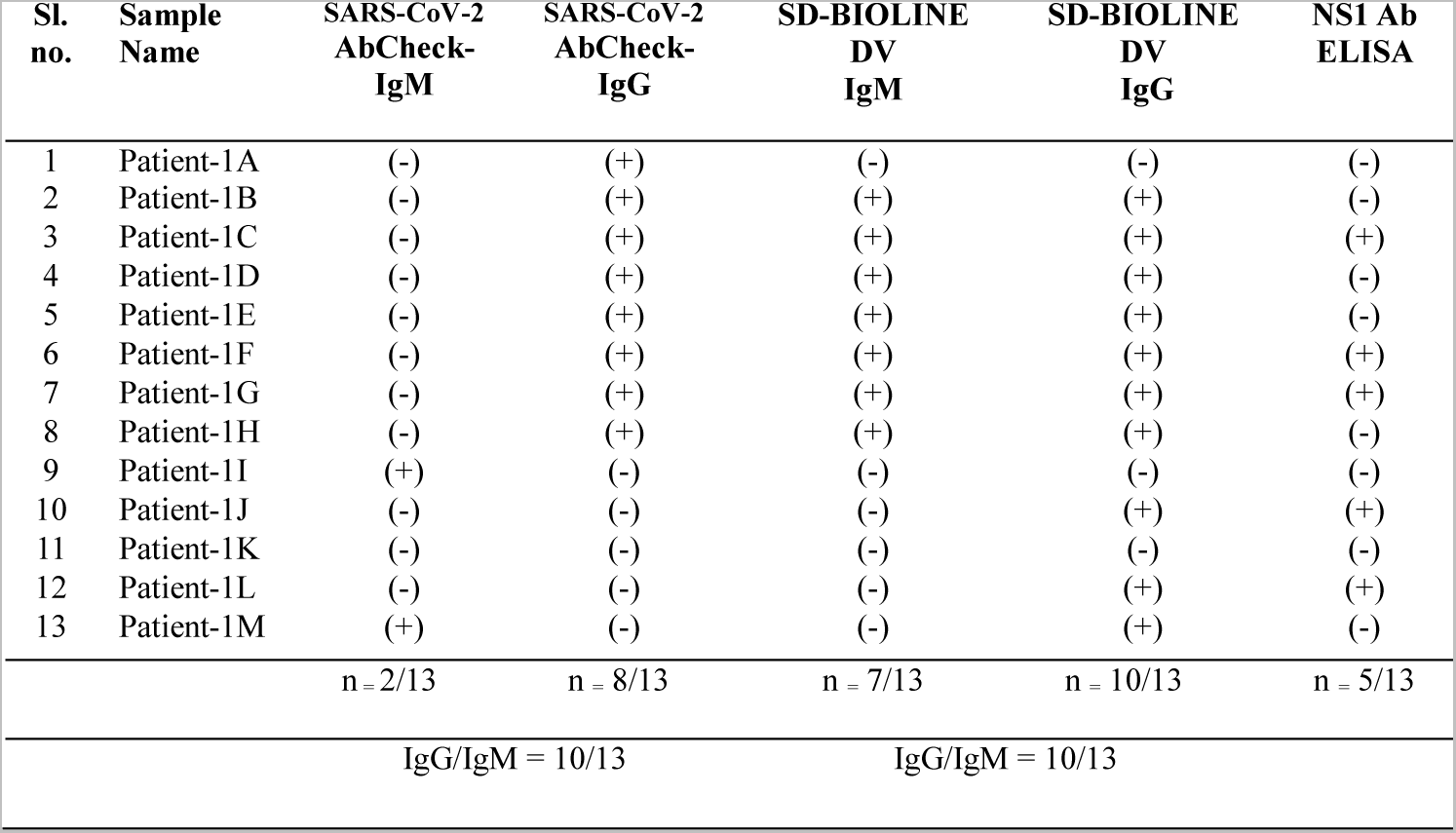
COVID-19 and Dengue strip test results of thirteen serum samples collected during January, 2022. The "+" sign signifies that the test result was positive; the "-" sign signifies that the test result was negative. Ab = antibody; Ag = antigen; DV = dengue virus; IgG = immunoglobulin G; IgM = immunoglobulin M; NS1 = nonstructural protein 1; SD-BIOLINE = Standard Diagnostics-Bioline; SL = serial.

#### 3.1.2. DV cross-reactivity status of COVID-19 serum samples collected during August, 2022

33 Omicron wave COVID-19 serum samples had been collected during August, 2022. In the SARS-CoV-2 Ab strip test, 25 of 33 (76%) samples were positive for only SARS-CoV-2 IgG (12 samples) or both (IgG, and IgM: 13 samples). In the DV Ab strip test, 11 of 33 serum samples (33%) were positive for DV IgG or IgG/IgM (10/33 DV IgG-positive; 1/33 IgG/IgM both positive). Only 8 of the 25 SARS-CoV-2 Ab-positive serum samples (32%) were cross-reactive in DV Ab LFIA and again 5 of these 8 samples (63%) had no evidence of past exposure to dengue (NS1 Ab-negative). All the samples were DV NS1 Ag negative (Table 2).

**Table 2.** COVID-19 and Dengue strip test results of thirty-three COVID-19 serum samples collected during August, 2022. The "+" sign signifies that the test result was positive; the "-" sign signifies that the test result was negative. Ab = antibody; Ag = antigen; DV = dengue virus; ELISA = enzyme-linked immunosorbent assay; IgG = immunoglobulin G; IgM = immunoglobulin M; NS1 = nonstructural protein 1; SD-BIOLINE = Standard Diagnostics-Bioline; SL = serial.

| Sl. No. | Sample Name | SARS-CoV-2<br>AbCheck-IgM | SARS-CoV-2<br>AbCheck-IgG | SD-BIOLINE<br>DV<br>IgM | SD-BIOLINE<br>DV<br>IgG | NS1 Ab<br>ELISA |
| --- | --- | --- | --- | --- | --- | --- |
| 1 | Patient -S01 | (+) | (+) | (-) | (+) | (+) |
| 2 | Patient -S02 | (+) | (+) | (-) | (+) | (-) |
| 3 | Patient -S03 | (+) | (+) | (-) | (-) | (-) |
| 4 | Patient -S04 | (-) | (+) | (-) | (+) | (-) |
| 5 | Patient -S05 | (+) | (+) | (-) | (+) | (-) |
| 6 | Patient -S06 | (+) | (+) | (-) | (-) | (-) |
| 7 | Patient -S07 | (-) | (+) | (-) | (-) | (-) |
| 8 | Patient -S08 | (-) | (+) | (-) | (-) | (-) |
| 9 | Patient -S09 | (-) | (+) | (-) | (-) | (-) |
| 10 | Patient -S10 | (+) | (+) | (-) | (-) | (-) |
| 11 | Patient -S11 | (+) | (+) | (-) | (-) | (-) |
| 12 | Patient -S12 | (-) | (+) | (-) | (-) | (-) |
| 13 | Patient -S13 | (+) | (+) | (-) | (-) | (-) |
| 14 | Patient -S14 | (-) | (+) | (-) | (-) | (-) |
| 15 | Patient -S15 | (+) | (+) | (+) | (+) | (+) |
| 16 | Patient -S16 | (-) | (+) | (-) | (+) | (-) |
| 17 | Patient -S17 | (-) | (+) | (-) | (-) | (-) |
| 18 | Patient -S18 | (-) | (-) | (-) | (+) | (+) |
| 19 | Patient -S19 | (-) | (-) | (-) | (-) | (-) |
| 20 | Patient -S20 | (-) | (-) | (-) | (-) | (-) |
| 21 | Patient -S21 | (-) | (+) | (-) | (+) | (+) |
| 22 | Patient -S22 | (-) | (+) | (-) | (-) | (-) |
| 23 | Patient -S23 | (-) | (+) | (-) | (-) | (-) |
| 24 | Patient -S24 | (+) | (+) | (-) | (-) | (-) |
| 25 | Patient -S25 | (+) | (+) | (-) | (+) | (-) |
| 26 | Patient -S26 | (-) | (+) | (-) | (-) | (-) |
| 27 | Patient -S27 | (-) | (-) | (-) | (+) | (+) |
| 28 | Patient -S28 | (-) | (-) | (-) | (+) | (+) |
| 29 | Patient -S29 | (+) | (+) | (-) | (-) | (-) |
| 30 | Patient -S30 | (+) | (+) | (-) | (-) | (-) |
| 31 | Patient -S31 | (-) | (-) | (-) | (-) | (-) |
| 32 | Patient -S32 | (-) | (-) | (-) | (-) | (-) |
| 33 | Patient -S33 | (-) | (-) | (-) | (-) | (-) |
|  |  | n = 13/33 | n = 25/33 | n = 1/33 | n = 11/33 | n = 6/33 |
|  |  | IgG/IgM= 25/33 |  | IgG/IgM=11/33 |  |  |

#### 3.1.3. DV cross-reactivity status of COVID-19 serum samples collected during September, 2022

Out of 9 serum samples collected in September, 2022, 6 samples (67 %) were SARS-CoV-2 IgG, IgM or both positive when tested in COVID-19 LFIA test. Only 2 of these 9 samples (22%) were DV IgG-positive. Among the 6 SARS-CoV-2 Ab-positive samples only one sample (P4) was DV IgG-positive. This must be due to cross-reactivity as this sample had no evidence of past dengue exposure (NS1 Ab-negative). Thus the DV cross-reactivity percentage was further reduced to 17 % (Table 3). As before, all the samples were tested for DV NS1 Ag and found negative (Table 3).

**Table 3:**
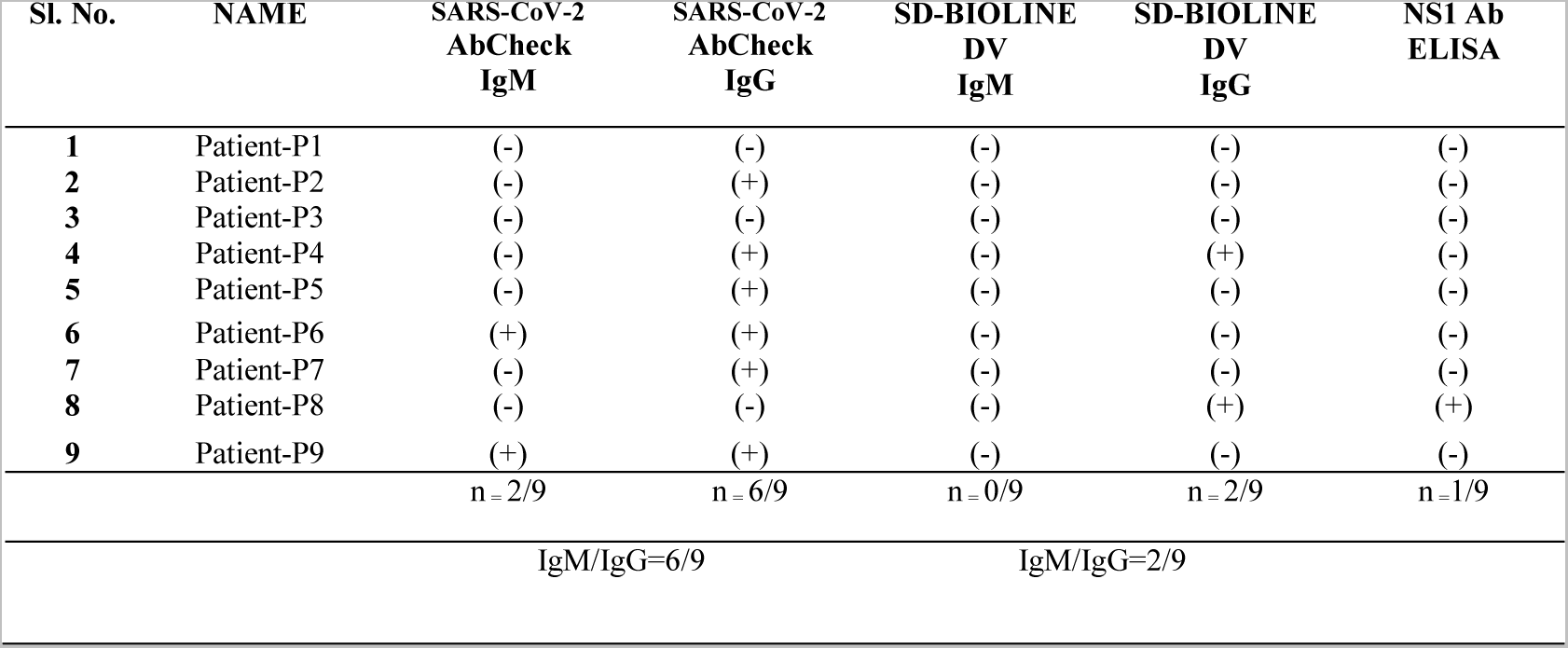
COVID-19 and Dengue strip test results of nine serum samples collected during September, 2022. The "+" sign signifies that the test result was positive; the "-" sign signifies that the test result was negative. Ab = antibody; Ag = antigen; DV = dengue virus; IgG = immunoglobulin G; IgM = immunoglobulin M; NS1 = nonstructural protein 1; SD-BIOLINE = Standard Diagnostics-Bioline; SL = serial

### 3.2. Virus neutralization assay using different DV serotypes and Omicron wave COVID-19 serum samples

All DV neutralization tests were carried out with Omicron wave COVID-19 serum samples, collected between January, 2022 and September 2022 (Table 4). DV1 neutralization was tested using 11 DV non-cross-reactive (CoV Ab+, DV Ab-, NS1 Ab-) COVID-19 serum samples (e.g. Patients-1A, S11, P2; Table 4, row 1, column 2) that highly reduced the viral titer in comparison to only DV1 or DV1 plus healthy serums (HS1-HS3) (Fig. 1a). As mentioned before, the HS1-HS3 pre-pandemic serum samples, collected from apparently healthy individuals, were free of DV Abs, SARS-CoV-2 Abs and DV RNA (RT-PCR) and DV NS1 (LFIA).

**Figure 1.**
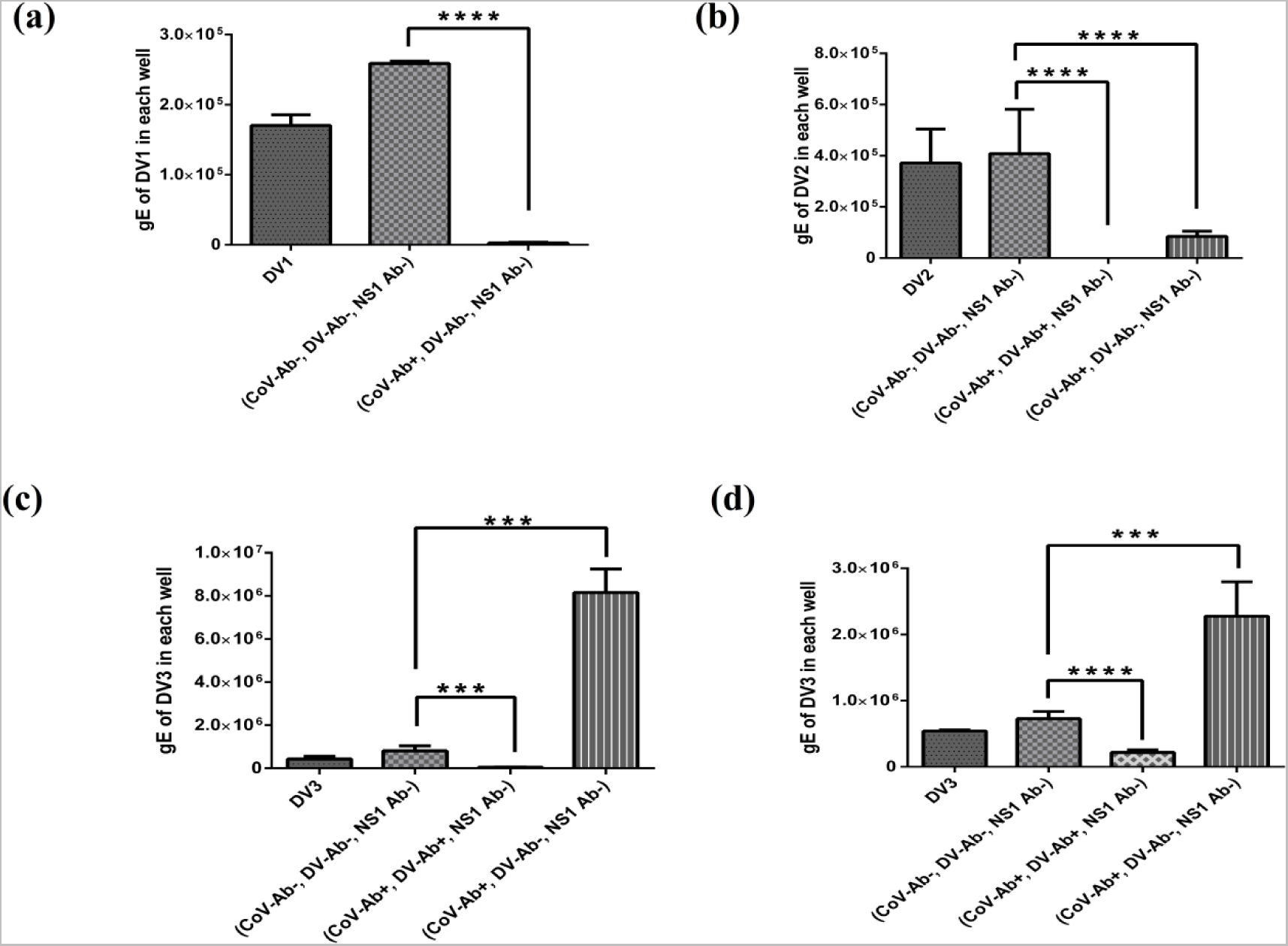
Result of DV (DV1, DV2 and DV3) neutralization assay using serum samples from Omicron wave SARS-CoV-2 infected patients. (a) DV non-cross-reactive (CoV Ab+, DV Ab-) serum samples decreased DV1 titres significantly (P<0.0001) as compared to healthy serum controls. (b) DV2 titres were reduced significantly by both DV cross-reactive (CoV Ab+, DV Ab+) (P<0.0001) and DV non-cross-reactive (P<0.0001) serum samples in comparison with healthy serums. (c and d) DV cross-reactive serum samples decreased the viral titres significantly [P=0.0002 for (c) and P<0.0001 for (d)], whereas DV non-cross-reactive serum samples showed a sharp rise [P=0.0003 for (c) and P=0.0008 for (d)] in DV3 titres (mean 10-fold as shown for the serum samples in (c) and 3-fold for the samples as shown in (d)) as compared to the healthy serum controls. Each column represents the mean virus titre and the error bar indicates SD. SD: standard deviation. gE: genome equivalent. Well: one well of a 96- well tissue culture plate.

**Table 4:** List of COVID-19 serum samples used in virus neutralization assay (VNT) against different DV serotypes.

| **DV serotypes** | **DV cross-reacting COVID-19 serums (CoV Ab+, DV Ab+) \*** | **DV non-cross-reacting COVID-19 serums (CoV Ab+, DV Ab-)** | **Pre-pandemic healthy serums (CoV Ab-, DV Ab-) (HS 1-3)** |
| --- | --- | --- | --- |
| DV1 | Neutralizing <sup>7</sup> | Neutralizing<br>[Patient 1A,1I,S06,S09,S11,S13,S22,S26,S29,P2,P7] | Non-neutralizing |
| DV2 | Neutralizing<br>[Patient-1B,1D,1E,1H,1M,S02,S04,S05,S16,S25,P4] | Neutralizing<br>[Patient 1A,1I,S06,S09,S11,S13,S22,S26,S29,P2,P7] | Non-neutralizing |
| DV3 | Neutralizing<br>[Patient-1B,1D,1E,1H,1M,S02,S04,S05,S16,S25,P4] | ADE<br>[Patient 1A,1I,S06,S09,S11,S13,S22,S26,S29,P2,P7] | Non-neutralizing |
| DV4 | Neutralizing<br>[Patient- 1B,1H,1M, S02, S05] | Neutralizing<br>[Patient 1A,S06,S09,S11,P2] | Non-neutralizing |

\*All the DV cross-reacting COVID-19 serums used in VNT were DV NS1 Ab-negative.

DV2 neutralization assay was done using 11 DV cross-reactive (e.g. Patients-1D, S04, P4; Table 4, row 2, column 1) and 11 DV non-cross-reactive (e.g. Patients-1A, S11, P2; Table 4, row 2, column 2) Omicron wave COVID-19 serum samples. All these samples greatly reduced the DV2 titres in comparison to only DV2 or DV2 plus Ab-negative healthy serum samples (Fig. 1b).

DV3 neutralization was tested using 11 DV cross-reactive and 11 DV non-cross-reactive COVID-19 serum samples. It was observed that the DV cross-reactive serum samples effectively neutralized DV3 (Fig. 1c, d). DV3 alone or DV3, incubated with healthy serums were used as controls. DV cross-reactive (SARS-CoV-2 Ab+, DV Ab+, NS1 Ab-) serum samples (e.g. Patients-S04, S16, S25; Table 4, row 3, column 1) neutralized DV3 and reduced the virus titres significantly compared to the Ab-negative serum controls. The above-mentioned 11 Omicron wave SARS-CoV-2 infected patients’ samples were DV cross-reactive (CoV-Ab+, DV-Ab+) but had no evidence of DV pre-exposure (i.e. DV RNA-; DV NS1- and DV NS1 Ab-). Both healthy serums and virus control showed a similar level of virus titre and the healthy serums were unable to neutralize DV3.

On the other hand, the Omicron wave SARS-CoV-2 infected patient samples (e.g. Patients-1A, S11, P2; Table 4, row 3; column 2) that were DV non-cross-reactive (CoV-Ab+, DV-Ab-, NS1 Ab-) significantly increased DV3 yields as compared to the healthy serums. For instance, serum from Patients-1A, S06, and S29 increased DV3 titres by approximately 10-fold compared to healthy serum controls (Fig. 1c) while serums from Patients-1I, S09, S11, S13, S22, S26, P2 and P7 significantly increased the virus titres by about 3-fold in comparison to the healthy control serums (Fig. 1d).

### 3.3. DV4 plaque neutralization assay using Omicron wave COVID-19 serum samples

The neutralization of DV4 plaques was carried out with representative Omicron wave serum samples (Table 4, row 4). Both the Omicron DV cross-reactive (Patients-1B, 1H, 1M, S02 and S05) and non-cross-reactive (Patients-1A, S06, S09, S11 and P2) COVID-19 serum samples prevented plaque formation. The wells that received only virus or virus incubated with healthy human serums (HS1, HS2, or HS3) showed visible plaques at 120h post-infection (Fig. 2).

**Figure 2.**
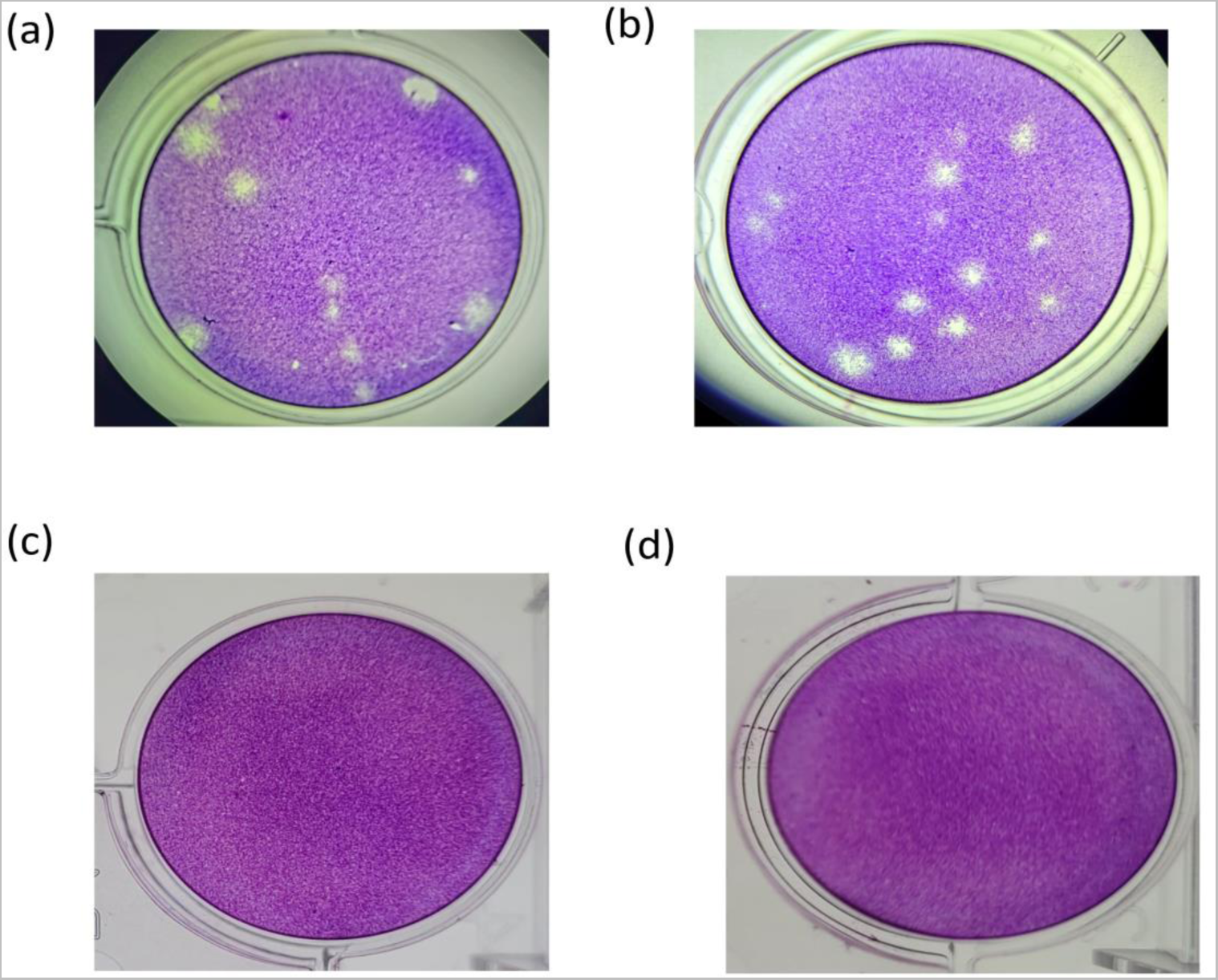
Representative image of the DV4 plaque neutralization assay using serum samples from Omicron wave SARS-CoV-2 infected patients. The wells which contained only DV4 (a) and DV4 incubated with healthy serum (b), showed 12 plaques on an average. The wells which contained DV4 along with DV cross-reactive (c) or non-cross-reactive (d) COVID-19 serum sample showed no plaque formation.

## 4. DISCUSSION

The salient findings of this study are summarized and discussed as follows:

a) With the advent and progression of the Omicron wave, the sensitivity of the SARS-CoV-2 Ab detection LFIA kit seemed to decrease. During the Delta wave, the sensitivity of the same LFIA kit used in this study, was around 90% (n=47/52)^7^. However, from January, 2022 to September, 2022, the same LFIA kit detected 77% SARs-CoV-2 Ab during January, 2022 (n=10/13); 76% during August, 2022 (n=25/33), and 67% during September, 2022 (n=6/9). This is perhaps due to the fact that the Omicron variants have approximately 30 mutations in the spike protein and several insertions/deletions compared to the wild-type^23^. So, the LFIA kit with wild-type SARS-CoV-2 spike antigen immobilized on it, may not detect the Abs to the Omicron variants as effectively as the earlier variants like Alpha, Beta, Gamma, or Delta. There seems to be another 10% drop in the overall sensitivity of the LFIA kit used (current sensitivity is approximately 80%) as the predominance of SARS-CoV-2 variants shifted from Delta (2020-21) to Omicron BA.1 (November, 2021 to March 2022)^24^ to mainly BA.2 in India (March, 2022 onwards)^25^ (Fig. 3).

**Figure 3.**
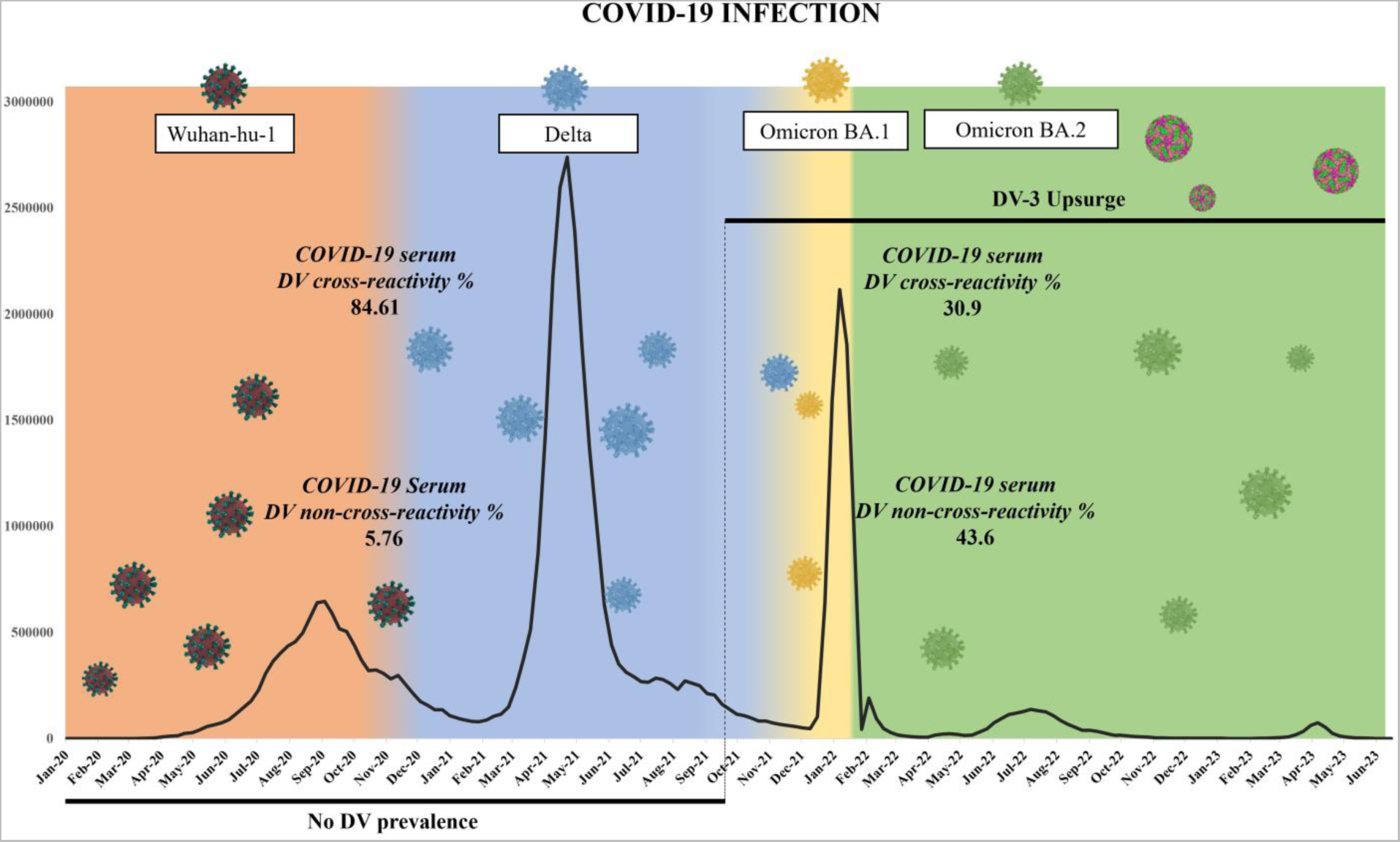
Month-wise pattern of COVID-19 infection waves (confirmed cases) in India and predominant variant(s) of SARS-CoV-2 from January, 2020 to June, 2023^25^ (Supplementary Table S5). From around October, 2020, the Delta variant wave predominated^26^. Omicron BA.1 variant was discovered for the first time in India around November, 2021. All of 2022 and the first half of 2023 are dominated by Omicron BA.2 variant^27^. The cross-reactivity percentage of COVID-19 serum with DV was 84.61% (n=44/52) in the first two waves of COVID-19^7^ which decreased to 30.9% (n=17/55) in the third wave of COVID-19 (this study). Also, there were negligible case reports of DV infection in Southeast Asia from January, 2020-October, 2021^28–30^. The DV3 strain became predominant in Southeast Asia from the latter half of 2021 and during 2022^18, 31, 32^. The upsurge of DV3 coincided with the decrease in the prevalence of DV cross-reactive COVID-19 and the dominance of BA.2 variant.

The UK Health Security Agency (UKHSA) designated BA.2 as a *Variant Under Investigation* (VUI) on January 19, 2022, due to its increasing dominance in many countries worldwide, including India, where it quickly replaced the Delta and Omicron BA.1 variants. UKHSA discovered that BA.2 had a higher growth rate than BA.1, with an “apparent growth advantage”^23, 33, 34^

b) Interestingly, with the introduction and evolution of the Omicron variants in India, the previously observed property of cross-reactivity of SARS-CoV-2 Ab-positive serum samples with DV in DV Ab detection LFIA test kit also decreased considerably. The cross-reactivity percentage declined from almost 80% in January, 2022 (n=8/10); 32% in August, 2022 (n=8/25); to roughly 17% (n=1/6) in September, 2022. The cross-reactivity between SARS-CoV-2 and DV was previously noted at 89% using the same DV Ab detection LFIA during the Delta wave^7^. This decreased cross-reactivity with DV was not unexpected because SARS-CoV-2 evolved in the presence of its own and DV Abs in dengue-endemic countries and the immune escape variants (eg. BA.2) are likely to evade both the selection pressures. This is in agreement with the fact that the Omicron variants were highly divergent variants and included some of the most critical mutations in the spike protein that were found to be associated with increased transmissibility and humoral immune escape potential^35–37^.

c) As mentioned previously, we and others have conclusively demonstrated that SARS-CoV-2 Abs that were DV cross-reactive could “cross-neutralize” DV1 or DV2^7, 10^. Here we further demonstrate that this type of SARS-CoV-2 cross-reactive Abs with no prior history of DV pre-exposure i.e. CoV spike Ab+; DV env Ab+; DV NS1 Ab-serum samples from COVID-19 patients could also neutralize DV3 (e.g. Patients-S04, S16 and S25) as well as DV4 (e.g. Patients-S02, S05). This observation perhaps provides one strong evidence as to why DV cases were reportedly decreased in India and many other dengue endemic regions like Indonesia, Sri Lanka and Guangzhou, China^3, 30^ during the COVID-19 Delta wave (2020-2021). During this time, the predominant SARS-CoV-2 Abs were DV cross-reactive^7^ and could “cross-neutralize” DV1-4.

d) As stated earlier, the DV cross-reactivity feature of SARS-CoV-2 Ab-positive serum samples showed a gradual decline with the progression of the Omicron wave and predominance of BA.2 variants. We monitored the DV cross-neutralizing status of this SARS-CoV-2 Ab+; DV env Ab-non-cross-reacting serum samples from COVID-19 patients collected between January-September, 2022. In this study, we have observed that this type of SARS-CoV-2 Abs could still cross-neutralize DV 1, 2 and 4 in standard virus neutralization tests (VNTs). This is further evidence that SARS-CoV-2 Abs even with non-detectable cross-reactivity with DV in LFIA (i.e. no detectable evidence of DV pre-exposure) could still neutralize DV in cell cultures, confirming and validating previous results^7, 10^. These serum samples were also DV RNA, NS1 Ag and NS1 Ab-negative.

However, they failed to “neutralize” DV3; instead caused “ADE” and resulted in increased DV3 replication and higher virus yields in Huh7 cell cultures, compared to controls (virus alone or virus incubated with normal human serum). This phenomenon was observed in DV3 VNTs on repeated occasions with serum samples (e.g. Patients-S06, S13, and S29) that were SARS-CoV-2 Ab-positive but DV env Ab-negative.

This type of COVID-19 serum (DV non-cross-reactive but SARS-CoV-2 Ab-positive) was observed at low level during the Delta wave in our earlier studies (n=3/47 i.e. 6%, collected between September 2020 and January 2021)^7^. Interestingly, these three serums didn’t neutralize DV1 in cell culture. The reason may lie in the spike backbone of those three variants found in low proportion during the Delta wave. Without matching sequence data, it is not possible to ascertain how these samples differed from BA.2, which was reported in India about one year later (after January 2022).

The proportion of DV non-cross-reactive but SARS-CoV-2 Ab-positive then increased during the Omicron wave to about 68% (n=17/25) during August, 2022. The preceding January, 2022 samples contained 20% (n=2/10) and the succeeding September, 2022 samples showed 83% (n=5/6) such serums but the overall sample size was too small to calculate percentage in either case. However, the trend was perceptible and this antibody feature was coincident with the predominance of the Omicron variants like BA.2. So, we believe that the Omicron variants, by virtue of increased mutations in spike protein elicited Abs that gradually decreased “detectable” cross-reactivity with DV in LFIA test. However, such Abs could still “cross-neutralize” DV serotypes 1, 2 and 4 but not DV3.

We acknowledge that we did not have matching spike protein sequence data to conclusively establish that the DV non-cross-reactive COVID-19 serum samples were exclusively elicited by Omicron variant BA.2 but the alignment of BA.2 predominance with the abundance of these serums in the population definitely points to a close association.

We had previously discussed that following the decline of DV cases during the active years of the pandemic (2020-21), there was a surge in DV3 cases observable from the late months of 2022 onwards in Southeast Asia including West Bengal, India, Bangladesh and Singapore^3, 38^. In the present times DV3 cases are being increasingly reported even from non-DV3 endemic countries like Brazil and Nigeria^15, 16, 39^.

It is a known fact that pre-existing Abs present in the serum from a primary DV infection, bind to the infecting DV particles during a subsequent infection with a different serotype and can lead to an overall increase in the viral replication. This phenomenon is known as antibody-dependent enhancement (ADE) ^40–42^. Again, ADE can also be observed between members of the same family of viruses. For example, Zika virus and dengue virus both belong to the *Flaviviridae* family and show cross-reactivity between their Abs^43, 44^. Many other studies also suggest that an initial infection with one virus might lead to the production of cross-reactive but non-neutralizing Abs that are capable of causing ADE in the subsequent infection with the other virus^45^. However, from literature survey, no experimental evidence has been found so far suggesting ADE between two different families of viruses. Our study shows for the first time that ADE of dengue virus, belonging to *Flaviviridae* family can result from pre-existing Abs to SARS-CoV-2, a member of the *Coronaviridae* family.

From all these evidences, we believe that the predominance of DV non-cross-reacting Omicron Abs is playing a key role behind the increased incidences of DV3 worldwide by means of DV3 ADE. It is still to be seen whether the emergent DV3 strains had other replicative advantages due to evolution in the presence of immune selection pressures due to co-evolving DV and SARS-CoV-2 in co-endemic regions.

It has been anecdotally observed that the DV3 cases during late 2022 were relatively severe in nature and caused increased hospitalizations and deaths even in young adults, features that match with ADE in humans. Over the past 50 years, DV2 has been identified as the most prevalent serotype in India^46^. Various studies have reported that Abs against DV can be detected in the serum samples even after several years from the onset of the infection^47, 48^. So, DV3 ADE is a natural expression due to pre-existing Abs to other DV serotypes (mainly DV2) because we know that non-cross-reactive serotype causes more severe disease^49^. Here, our study shows for the first time that additional ADE may occur due to SARS-CoV-2 Abs only in case of DV3 but not for the other three serotypes. SARS-CoV-2 Abs, studied so far, were all capable of neutralizing DV1, DV2 and DV4. Hence, greater precaution should be taken as DV3 surge and severity may be more in the upcoming years due to this “double ADE impact”, one due to non-neutralizing DV Abs and the other due to DV non-cross-reacting SARS-CoV-2 Abs.

## FUNDING

Funded by CSIR-India; grant number: MLP 130 (CSIR Digital Surveillance Vertical for COVID-19 mitigation in India). The grant was given to SB. The funders had no role in the study design, in the collection, analysis and interpretation of data; in the writing of the manuscript; and in the decision to submit the manuscript for publication.

## AUTHORS’ CONTRIBUTION

SB conceived and designed the study. SS, CD, AM, SD and CDC participated and designed the study and performed the experiments. Patient sample collection and SARS-CoV-2 RT-PCR testing was done by SG and his clinical collaborators. SS, CD, AM, SD and SB wrote the original draft of the manuscript. SB and AD performed the critical analysis of data. SB acquired the funding. The final manuscript was reviewed and edited by all the authors.

## ACKNOWLEDGEMENT

Authors acknowledge the support received from the Director of CSIR-IICB. SB also acknowledges AcSIR for support. SS, CD and AM acknowledge the support of CSIR for their CSIR Research Fellowship; SD acknowledges UGC for his Junior Research Fellowship.

## ETHICS STATEMENT

The study was conducted according to the guidelines of the Declaration of Helsinki, and approved by the Institutional Ethical Committees of Apollo Multispecialty Hospital, Kolkata and the Council of Scientific & Industrial Research-Indian Institute of Chemical Biology (CSIR-IICB), Kolkata.

## DECLARATION OF INTEREST

Nothing to declare.

## DATA AVAILABILITY

Further information and resource requests should be directed to and will be fulfilled by the corresponding author, Dr. Subhajit Biswas.

## Notes

### Competing Interest Statement

The authors have declared no competing interest.

