## Supplementary material for "Pre-dominance of dengue non-cross-reacting SARS-CoV-2 spike antibodies during the Omicron era and their role in the ADE-mediated surge of Dengue virus serotype 3": Table S1

**Table S1:** List of prevalent dengue virus serotypes in the pre- and post-COVID-19 era.

| Sl. No. | Location | Most prevalent serotype in respective years | | | | | |
|---|---|---|---|---|---|---|---|
| | | 2018 | 2019 | 2020 | 2021 | 2022 | 2023 |
| 1. | Eastern India (Kolkata) | DV-2 (87.14 %)[1] | DV-2 (70.54 %)[2] | Low DV cases[3] | DV-3 (48.3 %)[4] | DV-3 (46-52 %)[5] | NA |
| 2. | South India (Tamil Nadu, Kerala) | DV-2[6] | NA | NA | NA | DV-3[7] | DV-3[7] |
| 3. | Bangladesh | DV-2 (40.95 %)[8] | DV-3 (91.86%)[8] | Low DV cases[9] | DV-3 (87.5-100%)[8,10] | DV-3 (89%)[8,11] | NA |
| 4. | Sri Lanka | DV-2 (56%)[12] | DV-3 (47.62 %)[13] | Low DV-2 [13,14] | DV-3[13] | NA | NA |
| 5. | Brazil | DV-2 [15] | DV-1[15] | DV-2[15] | DV-1[15] | DV-1[15] | DV-3[16] |
| 6. | Singapore | DV-2[17] | DV-2[17] | DV-3[18] | DV-3[17] | DV-3[17] | DV-3[17] |
| 7. | Indonesia (Manado, North Sulawesi) | NA | DV-3[19] | Low DV[20] | NA | DV-3[21] | NA |
| 8. | China-Guangzhou | DV-1 (85%)[22] | NA | Low DV-2[23] | NA | NA | NA |
| 9. | Niger-Africa | NA | NA | NA | NA | DV-3[24] | NA |

*NA: No relevant data available

**Table S2:** Patient information on COVID-19 serum samples collected during January, 2022

| Sl. No. | Isolate name | Age range | Sex | Hospital ward | Approximate date of admission of the patient to the Hospital | Sample procurement date (8-9 days later) |
|---|---|---|---|---|---|---|
| 1 | Patient-1A | 51-55 | F | Standard ward | January, 2022 | January, 2022 |
| 2 | Patient-1B | 51-55 | F | Standard ward | January, 2022 | January, 2022 |
| 3 | Patient-1C | 51-55 | M | Standard ward | January, 2022 | January, 2022 |
| 4 | Patient-1D | 51-55 | M | Standard ward | January, 2022 | January, 2022 |
| 5 | Patient-1E | 56-60 | F | Standard ward | January, 2022 | January, 2022 |
| 6 | Parient-1F | 51-55 | F | Standard ward | January, 2022 | January, 2022 |
| 7 | Patient-1G | 56-60 | F | Standard ward | January, 2022 | January, 2022 |
| 8 | Patient-1H | 71-75 | M | Standard ward | January, 2022 | January, 2022 |
| 9 | Patient-1I | 66-70 | M | Standard ward | January, 2022 | January, 2022 |
| 10 | Patient-1J | 76-80 | F | Standard ward | January, 2022 | January, 2022 |
| 11 | Patient-1K | 81-85 | F | Standard ward | January, 2022 | January, 2022 |
| 12 | Patient-1L | 81-85 | F | Standard ward | January, 2022 | January, 2022 |
| 13 | Patient-1M | 41-45 | F | Standard ward | January, 2022 | January, 2022 |

**Table S3:** Patient information on COVID-19 serum samples collected during August, 2022

| Sl. No. | Isolate name | Age range | Sex | Hospital ward | Approximate date of admission of the patient to the Hospital | Sample procurement date (3-8 days later) |
|---|---|---|---|---|---|---|
| 1 | Patient-S01 | 61-65 | F | Intensive Care Unit | August, 2022 | August, 2022 |
| 2 | Patient-S02 | 76-80 | M | High Dependency Unit | August, 2022 | August, 2022 |
| 3 | Patient-S03 | 66-70 | M | Intensive Care Unit | August, 2022 | August, 2022 |
| 4 | Patient-S04 | 61-65 | M | Standard ward | August, 2022 | August, 2022 |
| 5 | Patient-S05 | 21-25 | F | Standard ward | August, 2022 | August, 2022 |
| 6 | Patient-S06 | 66-70 | M | High Dependency Unit | August, 2022 | August, 2022 |
| 7 | Patient-S07 | 76-80 | M | High Dependency Unit | August, 2022 | August, 2022 |
| 8 | Patient-S08 | 41-45 | M | High Dependency Unit | August, 2022 | August, 2022 |
| 9 | Patient-S09 | 46-50 | M | Standard ward | August, 2022 | August, 2022 |
| 10 | Patient-S10 | 71-75 | M | Intensive Care Unit | August, 2022 | August, 2022 |
| 11 | Patient-S11 | 66-70 | M | High Dependency Unit | August, 2022 | August, 2022 |
| 12 | Patient-S12 | 61-65 | F | Standard ward | August, 2022 | August, 2022 |
| 13 | Patient-S13 | 81-85 | M | Standard ward | August, 2022 | August, 2022 |
| 14 | Patient-S14 | 66-70 | M | High Dependency Unit | August, 2022 | August, 2022 |
| 15 | Patient-S15 | 86-90 | M | Standard ward | August, 2022 | August, 2022 |
| 16 | Patient-S16 | 66-70 | M | Standard ward | August, 2022 | August, 2022 |
| 17 | Patient-S17 | 71-75 | F | Intensive Care Unit | August, 2022 | August, 2022 |
| 18 | Patient-S18 | 61-65 | M | High Dependency Unit | August, 2022 | August, 2022 |
| 19 | Patient-S19 | 56-60 | M | High Dependency Unit | August, 2022 | August, 2022 |
| 20 | Patient-S20 | 61-65 | F | High Dependency Unit | August, 2022 | August, 2022 |
| 21 | Patient-S21 | 51-55 | M | Intensive Care Unit | August, 2022 | August, 2022 |
| 22 | Patient-S22 | 76-80 | F | Standard ward | August, 2022 | August, 2022 |
| 23 | Patient-S23 | 46-50 | M | Standard ward | August, 2022 | August, 2022 |
| 24 | Patient-S24 | 76-80 | M | Intensive Care Unit | August, 2022 | August, 2022 |
| 25 | Patient-S25 | 86-90 | F | Intensive Care Unit | August, 2022 | August, 2022 |
| 26 | Patient-S26 | 86-90 | M | Standard ward | August, 2022 | August, 2022 |
| 27 | Patient-S27 | 61-65 | F | High Dependency Unit | August, 2022 | August, 2022 |
| 28 | Patient-S28 | 76-80 | M | High Dependency Unit | August, 2022 | August, 2022 |
| 29 | Patient-S29 | 61-65 | F | Intensive Care Unit | August, 2022 | August, 2022 |
| 30 | Patient-S30 | 71-75 | F | High Dependency Unit | August, 2022 | August, 2022 |
| 31 | Patient-S31 | 76-80 | M | Intensive Care Unit | August, 2022 | August, 2022 |
| 32 | Patient-S32 | 46-50 | F | High Dependency Unit | August, 2022 | August, 2022 |
| 33 | Patient-S33 | 71-75 | F | Standard ward | August, 2022 | August, 2022 |

**Table S4:** Patient information on COVID-19 serum samples collected during September, 2022

| Sl. No. | Isolate name | Age range | Sex | Hospital ward | Approximate date of admission of the patient to the Hospital | Sample procurement date (9-10 days later) |
|---|---|---|---|---|---|---|
| 1 | Patient-P1 | 66-70 | F | Standard ward | September, 2022 | September, 2022 |
| 2 | Patient-P2 | 76-80 | M | Standard ward | September, 2022 | September, 2022 |
| 3 | Patient-P3 | 51-55 | F | Standard ward | September, 2022 | September, 2022 |
| 4 | Patient-P4 | 71-75 | M | Standard ward | September, 2022 | September, 2022 |
| 5 | Patient-P5 | 31-35 | M | Standard ward | September, 2022 | September, 2022 |
| 6 | Patient-P6 | 21-25 | F | Standard ward | September, 2022 | September, 2022 |
| 7 | Patient-P7 | 81-85 | F | Standard ward | September, 2022 | September, 2022 |
| 8 | Patient-P8 | 41-45 | F | Standard ward | September, 2022 | September, 2022 |
| 9 | Patient-P9 | 51-55 | M | Standard ward | September, 2022 | September, 2022 |

**Table S5:** Month-wise COVID-19 infection, from 2020-2023[25]

| Year | COVID-19 Infection |
|---|---|
| Jan-20 | 0 |
| Jan-20 | 2 |
| Feb-20 | 5 |
| Feb-20 | 0 |
| Feb-20 | 0 |
| Mar-20 | 32 |
| Mar-20 | 68 |
| Mar-20 | 253 |
| Mar-20 | 2395 |
| Apr-20 | 2587 |
| Apr-20 | 7356 |
| Apr-20 | 10,784 |
| Apr-20 | 13484 |
| May-20 | 23,959 |
| May-20 | 27,987 |
| May-20 | 40,941 |
| May-20 | 56,275 |
| Jun-20 | 64,484 |
| Jun-20 | 74,294 |
| Jun-20 | 89,539 |
| Jun-20 | 1,18,398 |
| Jul-20 | 1,76,388 |
| Jul-20 | 2,28,065 |
| Jul-20 | 3,07,904 |
| Jul-20 | 3,65,201 |
| Aug-20 | 4,02,287 |
| Aug-20 | 4,36,672 |
| Aug-20 | 4,55,258 |
| Aug-20 | 4,97,793 |
| Sep-20 | 6,40,545 |
| Sep-20 | 6,46,263 |
| Sep-20 | 5,91,913 |
| Sep-20 | 5,16,841 |
| Oct-20 | 5,04,433 |
| Oct-20 | 4,40,745 |
| Oct-20 | 3,70,260 |
| Oct-20 | 3,19,271 |
| Nov-20 | 3,23,672 |
| Nov-20 | 3,06,825 |

| | |
|---|---|
| Nov-20 | 2,81,227 |
| Nov-20 | 2,97,113 |
| Dec-20 | 2,12,807 |
| Dec-20 | 1,74,194 |
| Dec-20 | 1,56,627 |
| Dec-20 | 1,36,155 |
| Jan-21 | 1,36,319 |
| Jan-21 | 1,07,701 |
| Jan-21 | 96,548 |
| Jan-21 | 91,650 |
| Feb-21 | 80,180 |
| Feb-21 | 78,577 |
| Feb-21 | 86,711 |
| Feb-21 | 1,05,080 |
| Mar-21 | 1,14,068 |
| Mar-21 | 1,48,249 |
| Mar-21 | 2,40,082 |
| Mar-21 | 3,72,494 |
| Mar-21 | 5,13,885 |
| Apr-21 | 8,73,296 |
| Apr-21 | 14,29,304 |
| Apr-21 | 21,72,063 |
| Apr-21 | 25,97,285 |
| May-21 | 27,38,957 |
| May-21 | 23,87,663 |
| May-21 | 18,46,055 |
| May-21 | 13,64,668 |
| Jun-21 | 6,30,650 |
| Jun-21 | 4,41,976 |
| Jun-21 | 3,51,218 |
| Jun-21 | 3,12,250 |
| Jul-21 | 2,91,789 |
| Jul-21 | 2,68,843 |
| Jul-21 | 2,65,836 |
| Jul-21 | 2,83,923 |
| Aug-21 | 2,78,631 |
| Aug-21 | 2,58,121 |
| Aug-21 | 2,31,658 |
| Aug-21 | 2,70,796 |
| Sep-21 | 2,48,248 |
| Sep-21 | 2,11,242 |

| | |
|---|---|
| Sep-21 | 2,04,582 |
| Sep-21 | 1,61,158 |
| Oct-21 | 1,39,572 |
| Oct-21 | 1,14,244 |
| Oct-21 | 1,07,749 |
| Oct-21 | 97,832 |
| Nov-21 | 82,236 |
| Nov-21 | 81,771 |
| Nov-21 | 73,106 |
| Nov-21 | 67,110 |
| Dec-21 | 57,255 |
| Dec-21 | 49,765 |
| Dec-21 | 46,527 |
| Dec-21 | 1,02,330 |
| Jan-22 | 6,38,872 |
| Jan-22 | 15,94,160 |
| Jan-22 | 21,15,100 |
| Jan-22 | 18,55,258 |
| Feb-22 | 44,383 |
| Feb-22 | 1,91,052 |
| Feb-22 | 93,644 |
| Feb-22 | 46,836 |
| Mar-22 | 28,038 |
| Mar-22 | 16,850 |
| Mar-22 | 11,612 |
| Mar-22 | 8,672 |
| Apr-22 | 7,140 |
| Apr-22 | 6,826 |
| Apr-22 | 15,448 |
| Apr-22 | 21,643 |
| May-22 | 23,006 |
| May-22 | 19,405 |
| May-22 | 14,772 |
| May-22 | 16,672 |
| Jun-22 | 45,200 |
| Jun-22 | 74,675 |
| Jun-22 | 93,281 |
| Jun-22 | 1,12,456 |
| Jul-22 | 1,20,222 |
| Jul-22 | 1,27,948 |
| Jul-22 | 1,38,156 |

| | |
|---|---|
| Jul-22 | 1,31,056 |
| Aug-22 | 1,25,921 |
| Aug-22 | 1,07,732 |
| Aug-22 | 85,965 |
| Aug-22 | 68,703 |
| Sep-22 | 38,824 |
| Sep-22 | 38,829 |
| Sep-22 | 33,926 |
| Sep-22 | 26,373 |
| Oct-22 | 17,526 |
| Oct-22 | 16,815 |
| Oct-22 | 13,914 |
| Oct-22 | 9,524 |
| Nov-22 | 5,798 |
| Nov-22 | 2,638 |
| Nov-22 | 2,547 |
| Nov-22 | 1,830 |
| Dec-22 | 1,430 |
| Dec-22 | 1,130 |
| Dec-22 | 1,154 |
| Dec-22 | 1,543 |
| Jan-23 | 1,275 |
| Jan-23 | 1,116 |
| Jan-23 | 881 |
| Jan-23 | 758 |
| Feb-23 | 755 |
| Feb-23 | 799 |
| Feb-23 | 1,180 |
| Feb-23 | 1,803 |
| Mar-23 | 2,672 |
| Mar-23 | 4,928 |
| Mar-23 | 8,727 |
| Mar-23 | 18,458 |
| Apr-23 | 34,011 |
| Apr-23 | 61,499 |
| Apr-23 | 73,874 |
| Apr-23 | 53,400 |
| May-23 | 24,241 |
| May-23 | 11,044 |
| May-23 | 5,787 |
| May-23 | 3,283 |

| | |
|---|---|
| Jun-23 | 1,206 |
| Jun-23 | 692 |
| Jun-23 | 472 |
| Jun-23 | 145 |

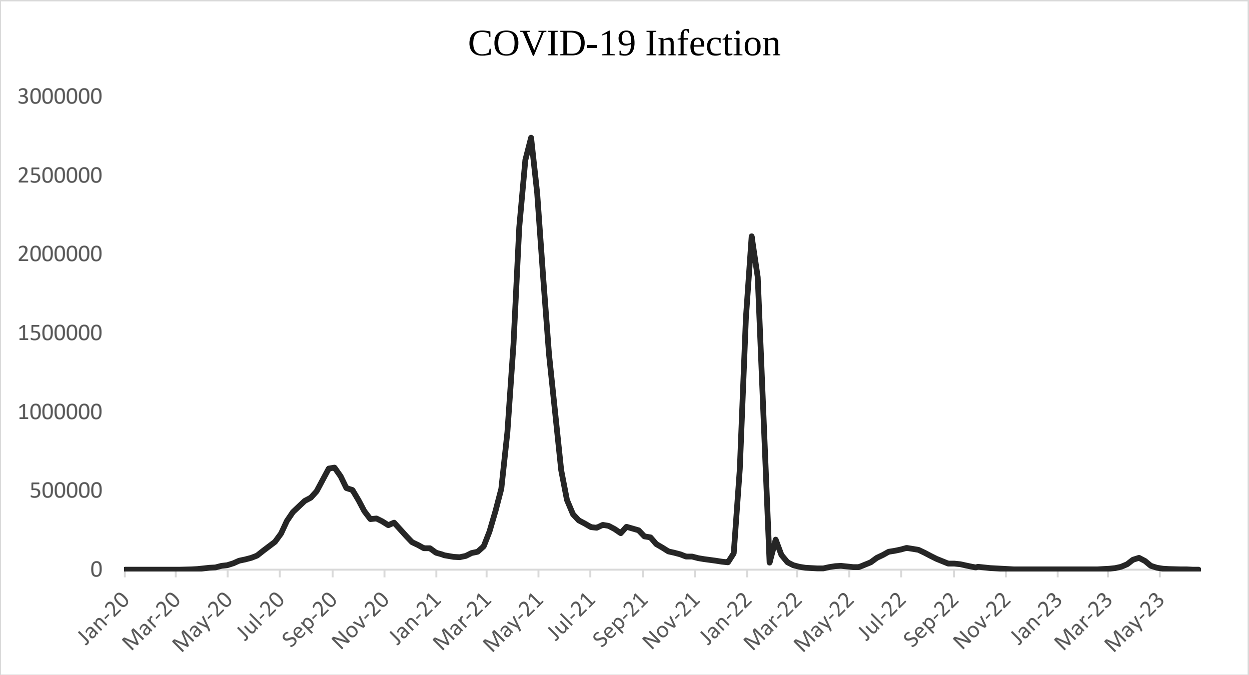

**Supplementary data**

**Table S1:** List of prevalent dengue virus serotypes in the pre and post-COVID-19 era.

**Table S2, S3 and S4:** Patient information of 55 COVID-1samples

**Table S5:** Month-wise COVID-19 infection, from 2020-2023
